# Assessing Population-level Target Product Profiles of Broadly-protective Human Influenza A Vaccines

**DOI:** 10.1101/2024.02.08.24302466

**Authors:** Qiqi Yang, Sang Woo Park, Chadi Saad-Roy, Isa Ahmad, Cécile Viboud, Nimalan Arinaminpathy, Bryan T. Grenfell

## Abstract

Influenza A has two main clades, with stronger cross-immunity to reinfection within than between clades. Here, we explore the implications of this heterogeneity for proposed cross-protective influenza vaccines that may offer broad, but not universal, protection. While the development goal for the breadth of human influenza A vaccine is to provide cross-clade protection, vaccines in current development stages may provide better protection against target clades than non-target clades. To evaluate vaccine formulation and strategies, we propose a novel perspective: a vaccine population-level target product profile (PTPP). Under this perspective, we use dynamical models to quantify the epidemiological impacts of future influenza A vaccines as a function of their properties. Our results show that the interplay of natural and vaccine-induced immunity could strongly affect seasonal clade dynamics. A broadly protective bivalent vaccine could lower the incidence of both clades and achieve elimination with sufficient vaccination coverage. However, a univalent vaccine at low vaccination rates could permit a resurgence of the non-target clade when the vaccine provides weaker immunity than natural infection. Moreover, as a proxy for pandemic simulation, we analyze the invasion of a variant that evades natural immunity. We find that a future vaccine providing sufficiently broad and long-lived cross-clade protection at a sufficiently high vaccination rate, could prevent pandemic emergence and lower the pandemic burden. This study highlights that as well as effectiveness, breadth and duration should be considered in epidemiologically informed TPPs for future human influenza A vaccines.

## 1 Introduction

Seasonal epidemics and occasional pandemics of human influenza A viruses cause substantial public health burden. Although vaccination is an important approach to mitigate this burden [3], current influenza A vaccines have significant limitations. First, they need to be evaluated annually for updates, due to the rapid turnover of antigenic variants. Second, existing vaccines can have low efficacy and narrow specificity (therefore cannot pre-emptively target potential pandemic influenza variants [14, 24, 15]). In addition, the frequent update of current vaccines may worsen vaccine hesitancy of individuals and increase the economic burden of vaccine purchases [2]. To tackle this challenge, there have been great efforts to develop a new generation of broadly-protective influenza vaccines [16, 22, 17, 1, 15]. However, there remain challenges in establishing broad protection. For example, a vaccine that targets one clade (e.g., clade 1) of human influenza A by raising stem-targeted antibodies might provide limited cross-protection to the other clade (e.g., clade 2) [17]. Widespread use of such a vaccine could have complex effects on influenza epidemiology where both clade 1 and 2 are already endemic, in ways that are not yet fully understood.

Here we construct a compartmental model to quantify vaccine impacts on seasonal influenza dynamics and potential pandemic invasions. The model incorporates host immunological history and clade co-existence. We model cross-clade interactions using H1N1 and H3N2 as respective representatives of H1 and H3 clades, choosing model parameters to match the broad behaviour of human influenza A in the USA in recent seasons. We assume the vaccine provides clade-level protection, i.e., strong cross-protection within one clade of influenza A but weaker cross-protection between clades. We focus on the impact of vaccinal immunity strength and duration on 1) seasonal strain dynamics of endemic infections and 2) immune escape at inter- and intra-clade levels, mimicking pandemic emergence. We can frame these variables by extending the standard notion of the Target Product Profile (TPP), which typically focuses on vaccine benefits to the individual, such as effectiveness. Although the TPP is an essential tool, we argue that its value for future influenza vaccines could be strengthened by considering the population-level processes (beyond vaccine effectiveness) more explicitly.

## 2 Methods

### 2.1 Mathematical model

When modelling human influenza strain dynamics, incorporating sufficient antigenic variation comes at the cost of complex host immune history. History-based models [27] take the host view, to track host infection history by different strains without including large variations in antigenicity. In contrast, while not including host immune history, status-based models [12] take the pathogen view, to track their impacts on hosts’ future infections by all other strains. To balance the complexity of antigenic variation and host immune history, we focus on the clade level of the influenza A virus phenotype and include relevant host immune history. We use *i* = 1 and *i* = 2 to represent the ’H1’ and ’H3’ clades, respectively. We construct our model (Figure 1), based on a previous 2-strain SIRS model on pertussis [19, 20] that incorporates host immune history. Our model allows immunity to reduce susceptibility against subsequent infections, and we refer to the proportional reduction in susceptibility as the *strength of cross-immunity* (using *θ* for infection-induced immunity, and *τ* for vaccine-induced immunity).

**Figure 1:**
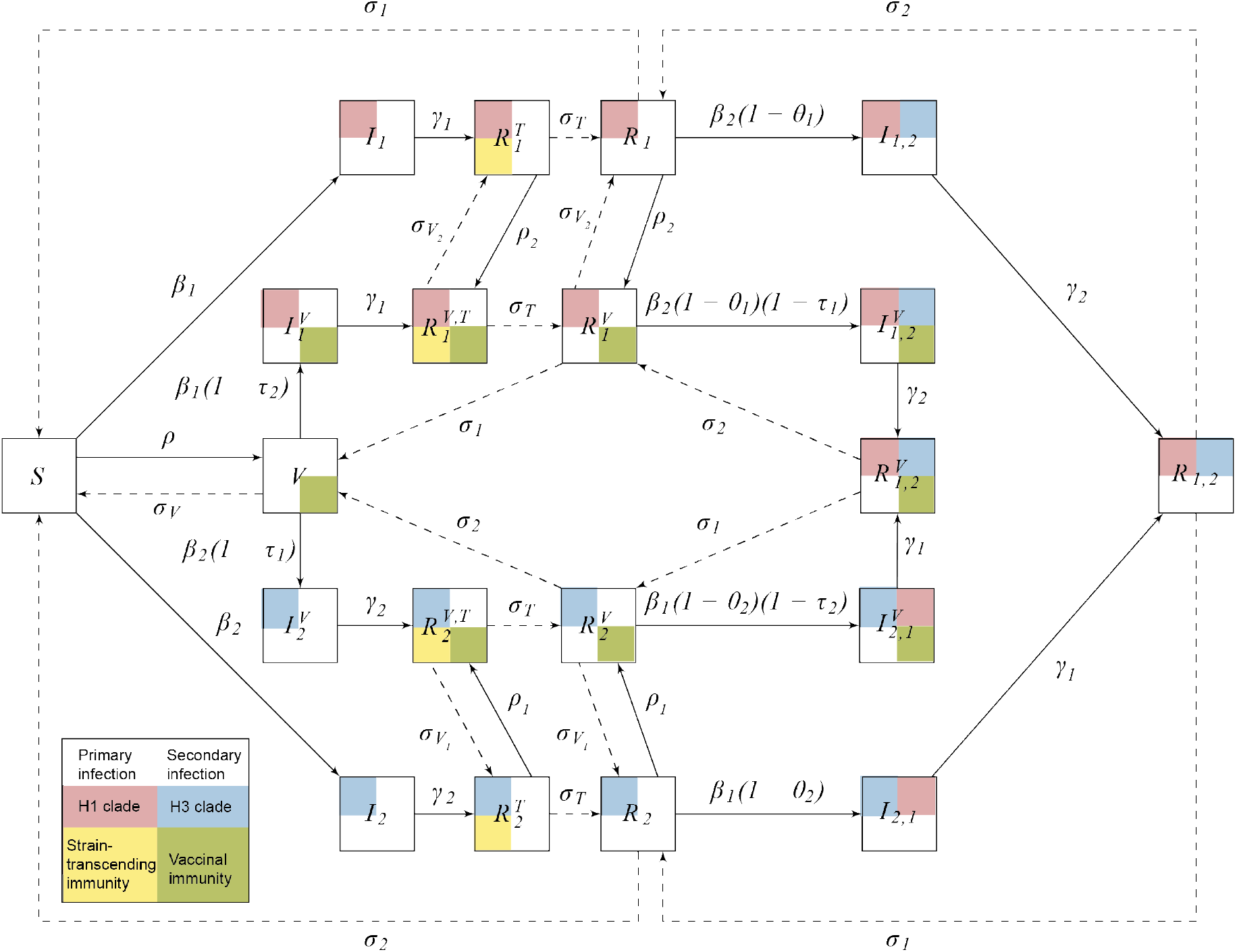
Model diagram. Births and deaths at rate *µ* per capita are not shown. The dashed lines show waning of immunity. The solid lines show infection, vaccination, or recovery. Each squared compartment can have up to four equal-sized ’sub-squares’ representing infection or immunity status. The left top sub-square represents primary infection; the right top is secondary infection. The infection is either of H1 clade (pink) or H3 clade (blue). The bottom two sub-squares denote strain-transcending immunity (yellow) and vaccinal immunity (green). Definitions of all compartments are in Table S1 and all parameters in Table S2. The ordinary differential equations of the model are in Supplementary S1.2

Hosts acquire immunity either via natural infection or vaccination. In the example of H1, we define transmission rate as *β*_1_. Following natural infection, strain-transcending immunity is induced first, i.e., short-term immunity against all clades, which can potentially shape the genetic diversity of seasonal human influenza [10]. This immunity wanes at rate *σ*_*T*_. After it wanes, the host can get infected by another clade with reduced susceptibility (*β*_2_(1 *− θ*_1_)); or immunity can further wane (at rate *σ*_1_), making the host fully susceptible to infections from both clades. After the host is infected with both clades, they are fully protected against all infections until immunity wanes. This immunity can wane (at rate *σ*_2_), making the host susceptible to the second clade again.

Hosts acquire vaccinal immunity via vaccination at rate *ρ*. This immunity wanes at rate *σ*_*V*_. We assume that vaccinal immunity can further enhance infection-induced immunity, and the resulting cross-immunity is modelled as a product of infection and vaccinal immunity. For example, a host recovered from H1 primary infection who is vaccinated has reduced susceptibility against H3 of *β*_2_(1 *− θ*_1_)(1 *− τ*_1_). We ignore the vaccine’s boosting effect on homotypic immunity when the host has already acquired ’perfect’ immunity (100% susceptibility reduction) against the clade via infection.

Regarding vaccine breadth, we model the following three vaccine scenarios:

- Bivalent vaccine. The vaccine reduces susceptibility against both clades proportionally by *τ*_1_ and *τ*_2_, respectively. The duration of vaccinal immunity is 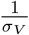 The vaccination rate is *ρ*_1_ = *ρ*_2_ = *ρ*. A bivalent vaccine is the ultimate goal of broadly-protective vaccine development; however, during initial development stages, it is likely that the vaccines would be univalent [17]:
- Univalent H1 vaccine. The vaccine fully reduces susceptibility against H1 clade (*τ*_2_ = 1) and partially against H3 clade (*τ*_1_ *<* 1). The duration of the immunity is 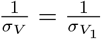 the vaccination rate is *ρ* = *ρ*_1_.
- Univalent H3 vaccine. The vaccine fully reduces susceptibility against H3 clade (*τ*_1_ = 1) and partially against H1 clade (*τ*_2_ *<* 1). The duration of the immunity is 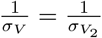. The vaccination rate is *ρ* = *ρ*_2_.

In each scenario, the strength and duration of the vaccinal immunity is same as that of infection-induced immunity 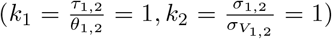 In further analysis, we vary *k*_1_ or *k*_2_ to be 0.5 or 2 to explore the vaccine’s impacts on seasonal epidemics (Figure 2 and 3).

**Figure 2:**
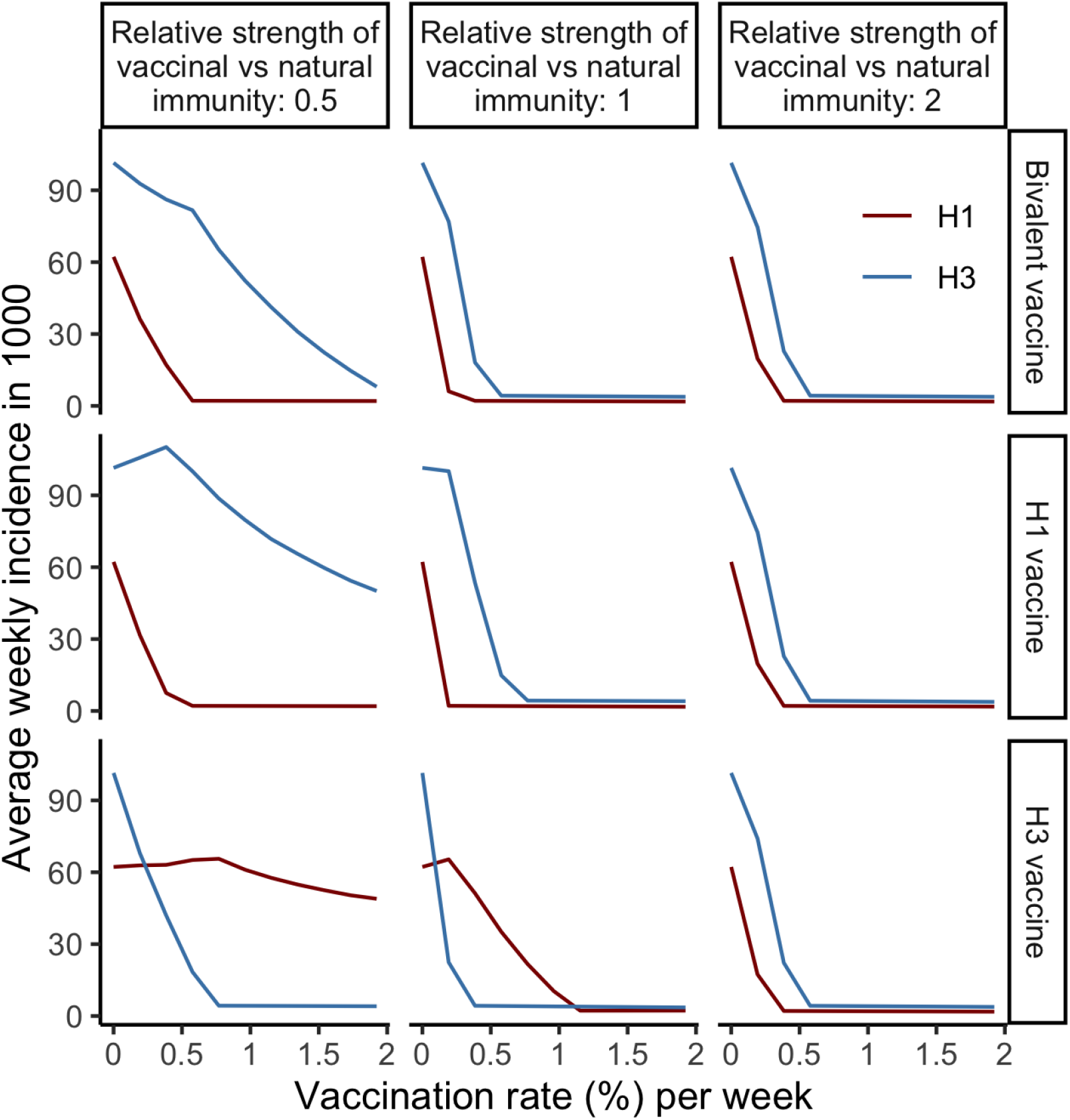
Average weekly incidence of H1 and H3 subtypes per 1000 population (*y* axis) vaccinated by vaccines with different target clades (rows) and immunity *strengths* (columns, defined as 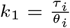), with changes of vaccination rate (%) per week (*x* axis). The results are calibrated with the best-fitting parameters (*θ*_1_ = 0.8, *θ*_2_ = 0.5, *a* = 0.04). Other parameter values are in Table S2

**Figure 3:**
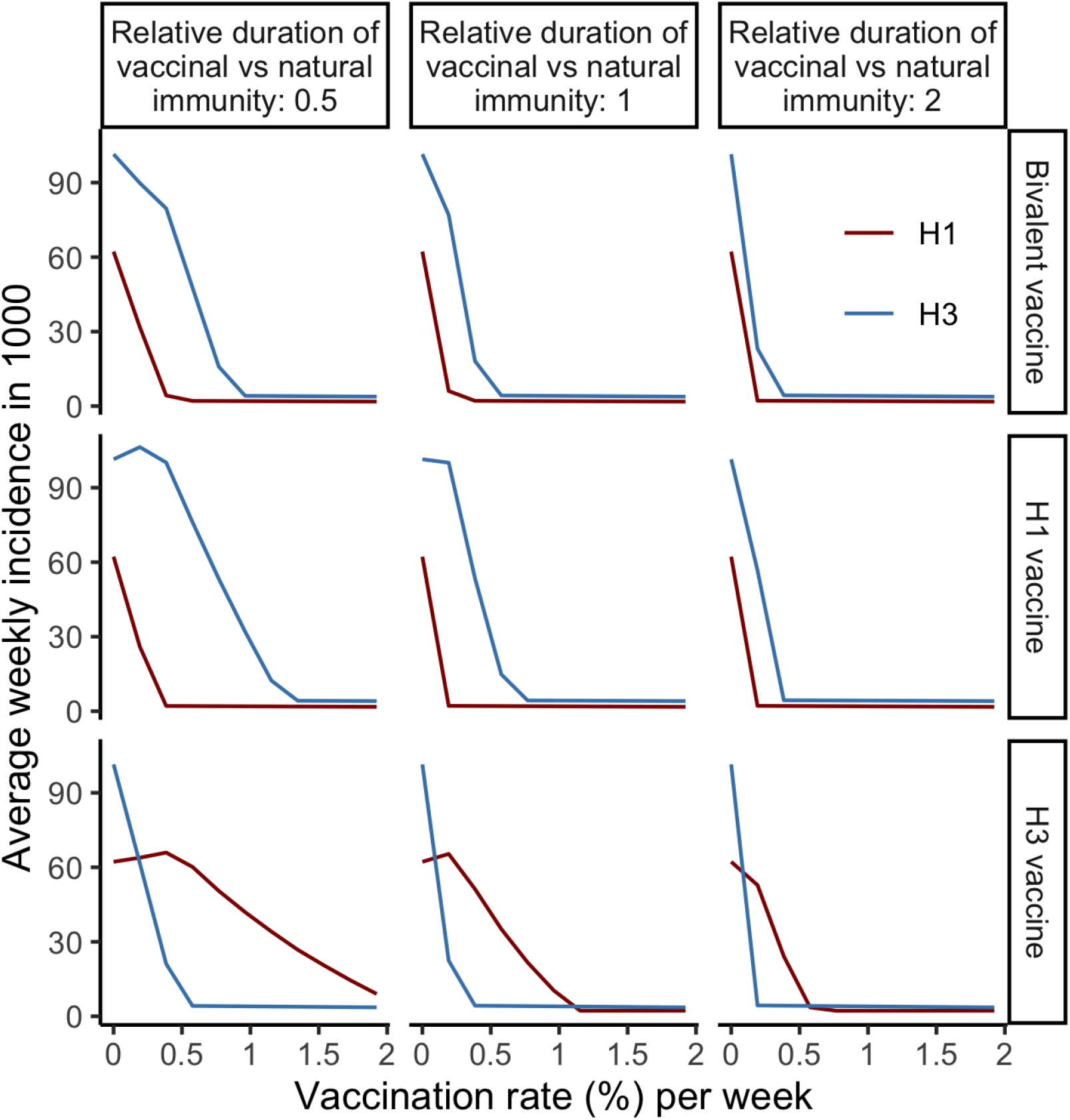
Average weekly incidence of H1 and H3 subtypes per 1000 population (*y* axis) vaccinated by vaccines with different target clades (rows) and immunity *durations* (columns, defined as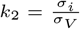), with changes of vaccination rate (%) per week (*x* axis). The parameters are same as in 2.

In addition, our model is *seasonally forced* because seasonality is a frequent characteristic of non-pandemic human influenza incidence [6]. The transmission rate *β*_1_(*t*) and *β*_2_(*t*) is determined by a standard sinusoidal function [9, 7]:

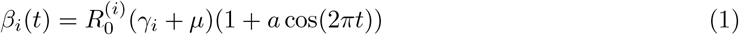

where *i* = 1, 2 represents H1 or H3 clade, 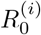 is the basic reproductive number of the clade *i, γ* is the recovery rate, *µ* is the birth rate and *a* is amplitude of the sinusoidal forcing. All parameters in the model and the sources of their values are listed in Table S2.

The model (Figure 1) is described by the ordinary differential equations in Supplementary materials S1.2. We set the initial conditions by assuming a 0.1% prevalence for H1 and H3 infections separately (therefore 0.2% in total). We assume that the remaining 99.8% of the population is fully susceptible. We then run deterministic simulations by numerically integrating the model for 100 years at semiweekly time steps to allow the system to reach its endemic phase. Subsequent analyses are based on this endemic phase reached from the initial condition. Sensitivity tests show that different initial conditions would have negligible effect on our results (Figure S1 and S2).

### 2.2 Model calibration

To ensure that the model captures subtype dynamics broadly consistent with that observed in temperate regions, we use influenza surveillance data during the 2004/2005-2018/2019 seasons in the USA. We use data on subtypes H1N1 and H3N2 to represent H1 and H3 clades. We omit the pandemic period (2008-2009, 2009-2010 and 2010-2011 seasons), shown as grey area in Figure S3. In order to capture broad features of influenza A epidemiology in the USA, we match model simulations to summary statistics of human influenza A incidence rates in the USA (Figure S3) as listed below: the auto-correlation and coefficient of variation of subtype incidence and cross-correlation between sub-type incidence. Key transmission parameters (Table S2) are drawn from a previous human influenza disease-dynamic modeling study focusing on Hong Kong [26].

The incidence rate (Figure S3) is estimated by combining influenza-like illness (ILI) surveillance data and laboratory-confirmed subtyping data as in the equation below, which is adapted from a previous study [8].

*Weekly incidence rate of subtype x = Outpatient ratio (Ratio of ILI patients among all hospital visits)* ^*\**^ *Ratio of influenza positive sample among all testings* ^*\**^ *Ratio of subtype x among all positive samples* where *x* =H1N1 or H3N2, and we used the ILI surveillance data weighted by the state population. Outpatient illness surveillance data and viral surveillance data were downloaded from FluView (US Centers for Disease Control and Prevention, CDC [11]). Viral surveillance data were collected from the US World Health Organization (WHO) Collaborating Laboratories and National Respiratory and Enteric Virus Surveillance System (NREVSS) laboratories [23].

Given the crudeness of the available data, we do not aim to capture detailed incidence in each season. Instead, we aim to match high-level patterns in the data, particularly in relation to the dominance patterns of the subtypes through time. We adjust values of three parameters: infection-induced cross-immunity strength (*θ*_1_ and *θ*_2_) and amplitude of sinusoidal seasonal forcing function (*a*) (to adjust the incidence). The incidence at each time step is calculated as the new infections with the focal clade regardless of the hosts’ immune statuses. The optimization metric is the sum of the absolute distance between simulation and data [11] of three summary statistics, including I) correlation between H1 and H3 seasonal incidence, II) coefficient of variation of H1 (and H3) seasonal incidence, III) auto-correlation of H1 (and H3) seasonal incidence. We run deterministic simulations by numerically integrating the model for 2000 years at semi-weekly time steps. We then calculated the statistics for every 12-year period (the first 800 years were removed as burn-in) and obtained a distribution of summary statistics. The optimization metric distribution across the parameter space (Figure S4) shows that when the seasonal forcing is weak, i.e., *a* = 0.04, the distances between simulation and data are the smallest, which is consistent with previous studies [9, 7]. For further analysis, we use the best parameter set *θ*_1_ = 0.8, *θ*_2_ = 0.5, *a* = 0.04 that gives the minimal distance across the parameter space (Figure S5). For sensitivity test, we choose a moderately-fit parameter set *θ*_1_ = 0.55, *θ*_2_ = 0.35, *a* = 0.04 that is more common across different *a* than other parameters (Figure S6). The results show that both parameter sets well match the dominance patterns of the subtypes in the data (Figure S7 and S8).

### 2.3 Invasion analysis of antigenic variants

To test the impact of the strength and duration of vaccinal immunity on pandemic emergence, we model a scenario where the population is exposed to (for simplicity) only one clade and is vaccinated against this clade. We simulate the model for 600 years at semi-weekly time steps to reach the endemicity of this clade. Then, a pandemic variant that fully escapes natural immunity is introduced to the population. We explore a range of basic reproduction numbers of the pandemic variant 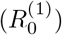 holding the endemic strain’s 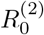 at 1.6, where i) 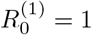, ii) 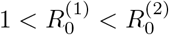, iii) 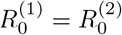, and iv)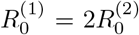. To test the vaccine impact, we vary the vaccination rate from 0 to 2% per week, the susceptibility reduction of the vaccine against the pandemic variant from 0 to 100% and vaccine immunity duration from 0.5, 1, 2, 4, 8 to 16 years. Using species invasion analysis similar to a previous study [20], we analyze the growth rate of the pandemic variant in the population as shown in the equation S7 (See Supplementary materials S1.3). We then numerically integrate the equations S1 to obtain deterministic dynamics of the system following the introduction of the pandemic variant with an incidence of one-millionth of the population size. The pandemic variant approximately follows exponential growth at the initial stage of invasion; therefore, we quantify the initial growth rate as the difference between the logarithm of the pandemic variant’s incidence on the third day and that on the second day, divided by the time difference (1 day). To quantify the persistence of the pandemic variant, we use trough depth, i.e., the minimal incidence of the pandemic variant after its initial peak.

## 3 Results

### 3.1 Impacts on seasonal epidemics of vaccinal immunity breadth, strength and duration

For each vaccinal immunity breadth scenario (see Methods), we explore a range of different strengths (Figure 2) and durations of vaccine-induced cross-protection (Figure 3) relative to infection-induced immunity. The robustness of the results is confirmed using not only the best-fitting parameters set, but also alternative, moderately-fitting parameter sets (Figure S9 and S10).

A bivalent vaccine (top row in Figure 2 and 3) would eliminate both clades without the potential of an incidence increase. The vaccination rates required to eliminate H1 and H3 are lower when the vaccinal immunity is stronger or longer. Due to higher *R*_0_, the H3 clade requires a higher vaccination rate to eliminate. When the vaccine targets one clade (either H1 or H3 clade, bottom two rows in Figure 2), providing the same or weaker immunity than natural infection (the left and middle columns in Figure 2), low vaccination rates could lead to an increase in the incidence of the non-target clade. The peak of the incidence occurs at the vaccination rate that eliminates the target clade. In contrast, when the vaccine-induced cross-immunity is stronger than natural immunity, the incidence of the off-target clade always decreases with the vaccination rate (the right column in Figure 2). The reason for this contrast lies in the interplay of natural and vaccinal cross-immunity:

- When the vaccination rate is 0, the cross-immunity is completely infection-induced.
- Before the target clade is eliminated, the cross-immunity comprises vaccinal and natural immunity that is stronger than vaccinal immunity alone.
- When the target clade is eliminated, the cross-immunity against the non-target clade is completely vaccine-induced.

Therefore, depending on the relative strength of vaccinal and natural immunity, the combined strength of vaccinal and natural cross-immunity becomes either stronger or weaker with the increase in vaccination rate.

Similarly, when the strength of vaccinal immunity is the same as infection-induced immunity, the H1 or H3 vaccine could eliminate the target clade at a low vaccination rate (bottom two rows in Figure 3). However, the incidence of the non-target clade could increase slightly at low vaccination rates. The shorter the duration of vaccine immunity compared to the infection immunity, the higher the minimum vaccination rate required to prevent any incidence increase of non-target clade. Additionally, when vaccine-induced immunity duration is twice as long as infection-induced immunity, the vaccination rate required for eliminating one clade and reaching the incidence plateau of the other clade is correspondingly reduced to 50% (Figure 3).

### 3.2 Impacts on pandemic emergence and persistence of vaccinal immunity breadth, strength and duration

*How might vaccinal cross-immunity strength and duration collectively impact the emergence of a pandemic variant in a vaccinated population?* As a simple representation of pandemic emergence, we assume the population is exposed to only one endemic clade, e.g., H3 clade (i.e., *i* = 2), and is vaccinated by the vaccine targeting H3 clade. Then it is exposed to a pandemic variant. We made a pessimistic assumption that H3 infection does not induce cross-immunity against the pandemic variant. The analytical result (Supplementary materials S1.3) shows that even when the pandemic variant has a lower basic reproductive number than the endemic strain, i.e., 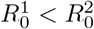, the pandemic can still emerge in the population, depending on the vaccine characteristics. Results in Figure 4 show how the initial growth rate of the pandemic variant depends on vaccine characteristics, including vaccination rate and immunity strength and duration. First, the boundary of pandemic emergence (shown with the red curve) moves towards less susceptibility reduction by the vaccine, for lower values of 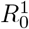 (Figure 4). In line with intuition, this indicates that the threshold of vaccine cross-immunity strength to prevent the pandemic is strongly dependent on the basic reproductive number of the pandemic variant. In addition, longer vaccine-induced cross-immunity duration lowers the vaccination rate threshold and vaccinal immunity strength level for preventing pandemic emergence. Following introduction, the pandemic variant can either 1) fail to emerge, 2) emerge and co-exist with the endemic strain, or 3) emerge and eliminate the endemic strain. Under limited vaccinal protection, the endemic and pandemic strains co-exist (corresponding colored areas in Figure 5 and S11 under the same vaccine characteristics). Vaccination characteristics and fitness of the pandemic strain decide the emergence of the pandemic strain (Figure 5 and S12). Any of the following vaccine characteristics could more easily prevent the emergence of the pandemic variant: higher vaccination rate, longer immunity duration and more susceptibility reduction against the pandemic variant (Figure 5). Interestingly, at the same vaccination rate, more susceptibility reduction could increase the incidence during the trough following invasion (Figure S12). This could be because stronger cross-immunity against the pandemic variant allows slower spread of the variant at the beginning. Due to the demographic stochasticity, when the trough depth is lower than 10^−^6, the pandemic variant could still become extinct (e.g., Figure S12) in a very large city. Therefore, we define the pandemic persistence threshold (orange curve in Figure 5) as 10^−^6. Additionally, the result suggests that the pandemic invasion trough depth might have a non-monotonic relationship with vaccine strength and vaccination rate. This non-monotonic relationship on persistence has been shown in other studies [18].

**Figure 4:**
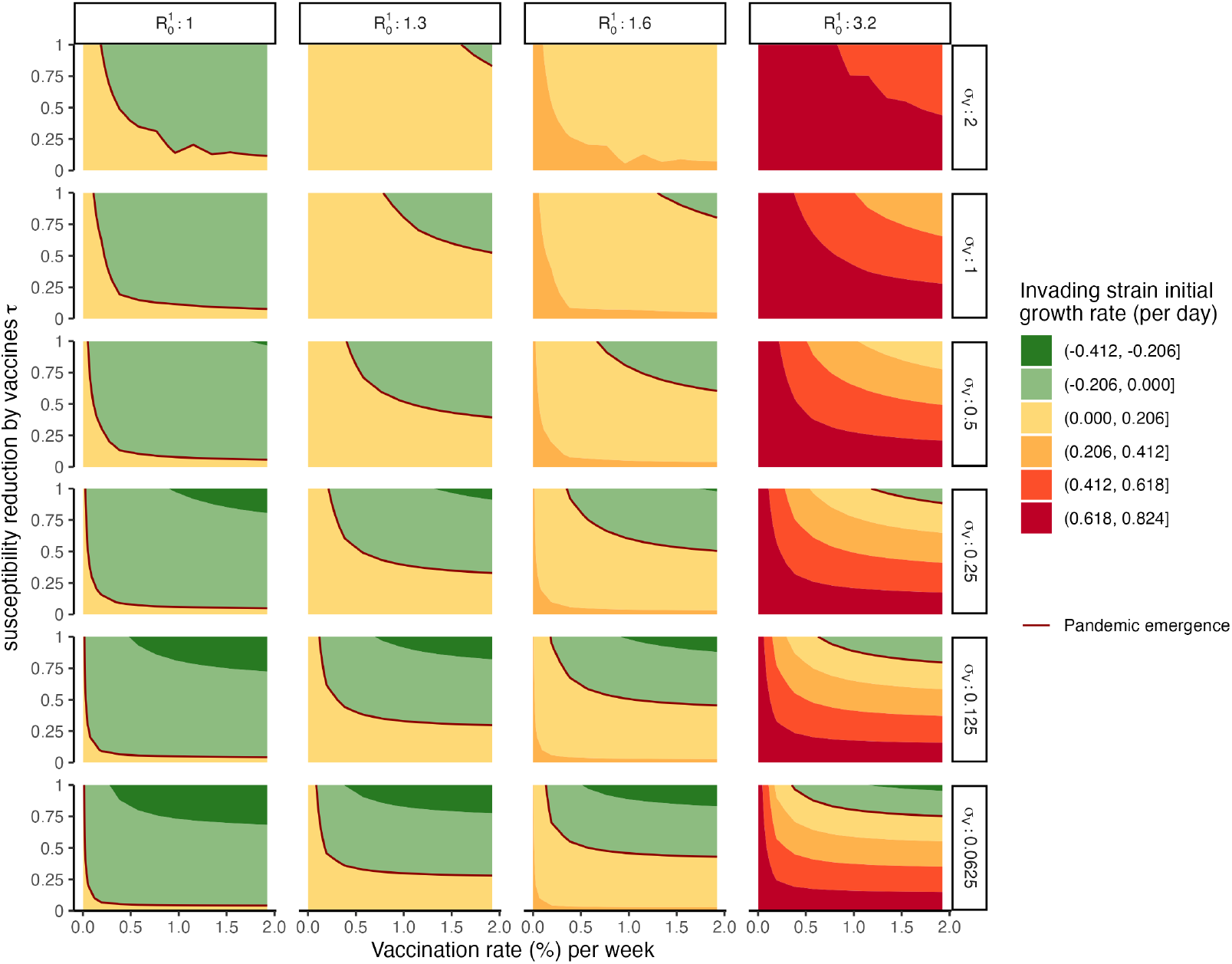
Emergence of a pandemic variant with the basic reproduction number 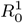 (column), which is measured by initial growth rate coloured by greens (failure of emergence) and yellow-to-reds (emergence), under vaccination of different immunity strength (*y* axis), duration (row) and vaccination rate *ρ* (%) per week (*x* axis). In these simulations, the pandemic variant completely escapes natural cross-immunity (*θ*_2_ = 0), the average infection-acquired immunity period is 2.7 years, the average infectious period is 3.03 days, and the average life span is 75 years. The initial growth rate is defined as the incidence growth of the pandemic variant between the second and third day of the invasion.

**Figure 5:**
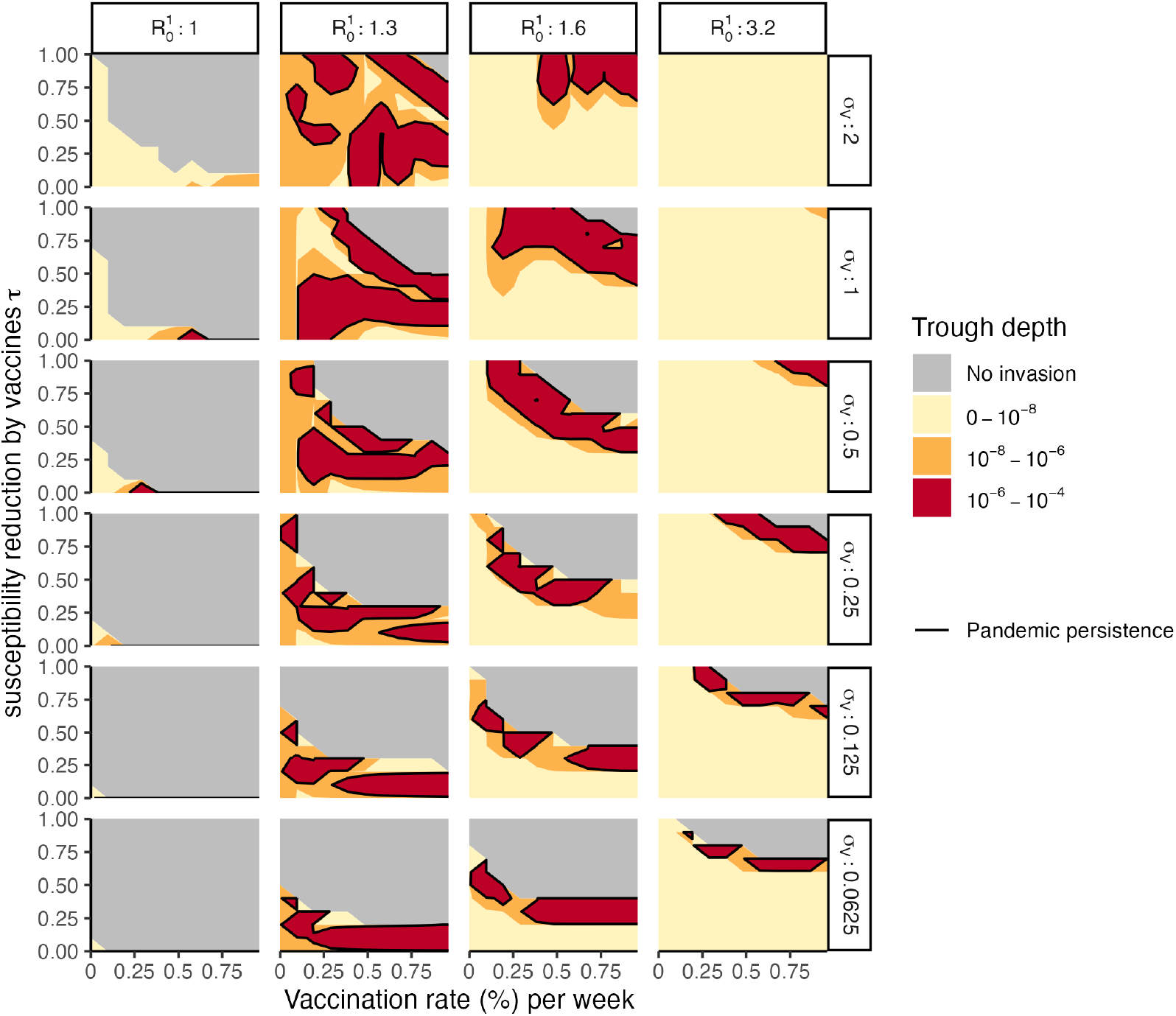
Persistence of the pandemic variant 5 years after its emergence, measured by trough depth of its incidence ((the minimal incidence), under vaccination of different immunity strength (*y* axis), duration (row) and vaccination rate *ρ* (%) per week (*x* axis), with different basic reproduction number 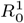 (column). The grey area is when the pandemic variant fails to emerge. The black curves indicate the transition between persistence and failure in the emergence of the pandemic variant. Other parameters are same as those in Figure 4.

## 4 Discussion

While TPPs of a vaccine are rightly informed by individual-level factors such as safety and efficacy against a symptomatic endpoint, the population-level epidemiological impacts of vaccines are less considered. By quantifying the potential epidemiological impacts, our work illustrates how TPPs could be complemented with population-level effectiveness, breadth, and immunity duration. What indicators would be important in such population-level TPPs? Based on our analysis, Table 1 lists some important characteristics, as well as some that should be addressed in future studies.

**Table 1:** Population-level Target Product Profiles (PTPPs) for broadly-protective human influenza A vaccines.

| Vaccine characteristic | Preferred | Study sources |
| --- | --- | --- |
| Breadth | Complementary vaccines for all clades (in the absence of universal protection) | This study |
| Strength* | No weaker than infection-induced immunity | This study |
| Duration* | No shorter than infection-induced immunity | This study |
| Immunity type | Susceptibility-reduction (preferred to infectiousness reduction) | This study and [5] |
| Dose regimen | Annual vaccination | [21] |
| Vaccination rate | High rate (the sufficient rate depends on the vaccination strategy) | This study |
| Vaccination acceptability and achievable coverage | / | Future studies |
| Interaction of vaccine immunity with prior immunity | / | Future studies |
| Evolutionary impacts on seasonal influenza viruses | / | Future studies building on existing literature ([4]) |

\* In practice, the strength and duration of the vaccine are usually correlated [14].

Specifically, our results show how the interplay between natural and vaccine-induced cross-immunity affects both clade dynamics and pandemic emergence. In this process, both vaccine immunity strength and cross-immunity strength are important. Notably, when a broadly protective vaccine provides limited protection against a non-target clade, this clade might cause a larger epidemic at low vaccination rates, than in the absence of vaccination. In contrast, the bivalent vaccine scenario never permits a larger epidemic and could eliminate both clades at similar vaccination rates. Therefore, our results highlight that adopting vaccines for against both clades would be essential for the broadly-protective vaccine deployments.

Additionally, when a vaccine-escape variant emerges, the population might be more vulnerable to the variant [5] than in the absence of the vaccine. For example, in theory, a vaccine providing cross-immunity by reducing infectiousness would reduce transmission at the population level and improve herd immunity in the long term [4]; however, such a vaccine might permit larger pandemics when the coverage is low [5], by increasing the population’s vulnerability to a pandemic invasion by a non-target variant. These existing studies focused on hypothesized vaccines that only reduce the infectiousness without reducing the susceptibility to infection, consistent with the effect of non-sterilizing T-cell immunity. Currently, promising vaccine candidates are being developed to reduce susceptibility to infection, e.g., one targeting the HA-stalk region [17]. Our results illustrate the potential of the epidemiological impacts of such vaccines: they might have a lower risk of permitting a larger pandemic than those that reduce infectiousness.

However, due to the lack of relevant experimental data on the mechanisms of vaccinal immunity, our results remain theoretical. Other caveats of this work include that the model assumes a homogeneous infection in a well-mixed population; for example, age or spatial structure of hosts or antigenic imprinting [13] could favor the invasion of the pandemic variant. The model also does not incorporate subtype diversity within clades of influenza viruses, nor does it address the effect of immunity on severity. In addition, we did not assess potential evolutionary impacts of the broadly-protective vaccines [4]. Furthermore, our model calibration only matches the subtype dominance pattern. The model could be refined by fitting time series of influenza incidence in the future, if more refined data become available. Finally, we assume that vaccine-induced and infection-induced immunity are independent, but in practice, they might oppose each other due to a possible ceiling effect, i.e., the level of protection cannot exceed a threshold. Nonetheless, even our simple epidemiological model provides insights into the potential value of population-level TPPs: the importance of considering immunity breadth, strength and duration in broadly-protective vaccine design and deployments. To advance understanding of population-level TPPs, future studies could incorporate virus genomic data and human immune data by applying immuno-epidemiological phylodynamic modelling approaches. These advances depend crucially on maintaining and enhancing epidemiological, genomic, and immune surveillance against influenza and other imperfectly-immunizing pathogens.

## Data Availability

All data produced are available online at

https://github.com/kikiyang/BroadFluAVacModel

## 5 Acknowledgements

This work was funded by FluLab. Q.Y. acknowledges funding from the Center for Health and Wellbeing, Princeton University, via Graduate Funding for Health-Focused Research. S.W.P. acknowledges funding from Princeton University via a Charlotte Elizabeth Procter Fellowship. C.M.S.-R. acknowledges funding from the Miller Institute for Basic Research in Science of UC Berkeley via a Miller Research Fellowship. B.T.G acknowledges support from Princeton Catalysis Initiative. The authors thank Bin Zhang for suggesting potential datasets. Disclaimer: The findings and conclusions in this report are those of the authors and do not necessarily represent the official position of the US National Institutes of Health or Department of Health and Human Services.

## 6 Author contributions

N.A. and B.T.G. contributed to funding acquisition and supervision. Q.Y., S.W.P., C.M.S.-R., N.A., and B.T.G. contributed to model conceptualization. Q.Y. and S.W.P. contributed to formal analysis and visualization. Q.Y., C.V., C.M.S.-R. and I.A. contributed to data curation. Q.Y., N.A. and B.T.G. wrote the original draft. All authors contributed to reviewing and editing the draft.

## S1 Supplementary information

The supplementary information includes:

- Dataset and code availability
- Ordinary differential equations of the model, Supplementary Tables S1-2
- Invasion analysis of a pandemic variant
- Supplementary Figures S1-11

### S1.1 Dataset and codes

See R codes and data on our GitHub repository: https://github.com/kikiyang/broadFluVacModel

### S1.2 Ordinary differential equations of the model

See Table S1 and S2 below for a description of all symbols in the model.

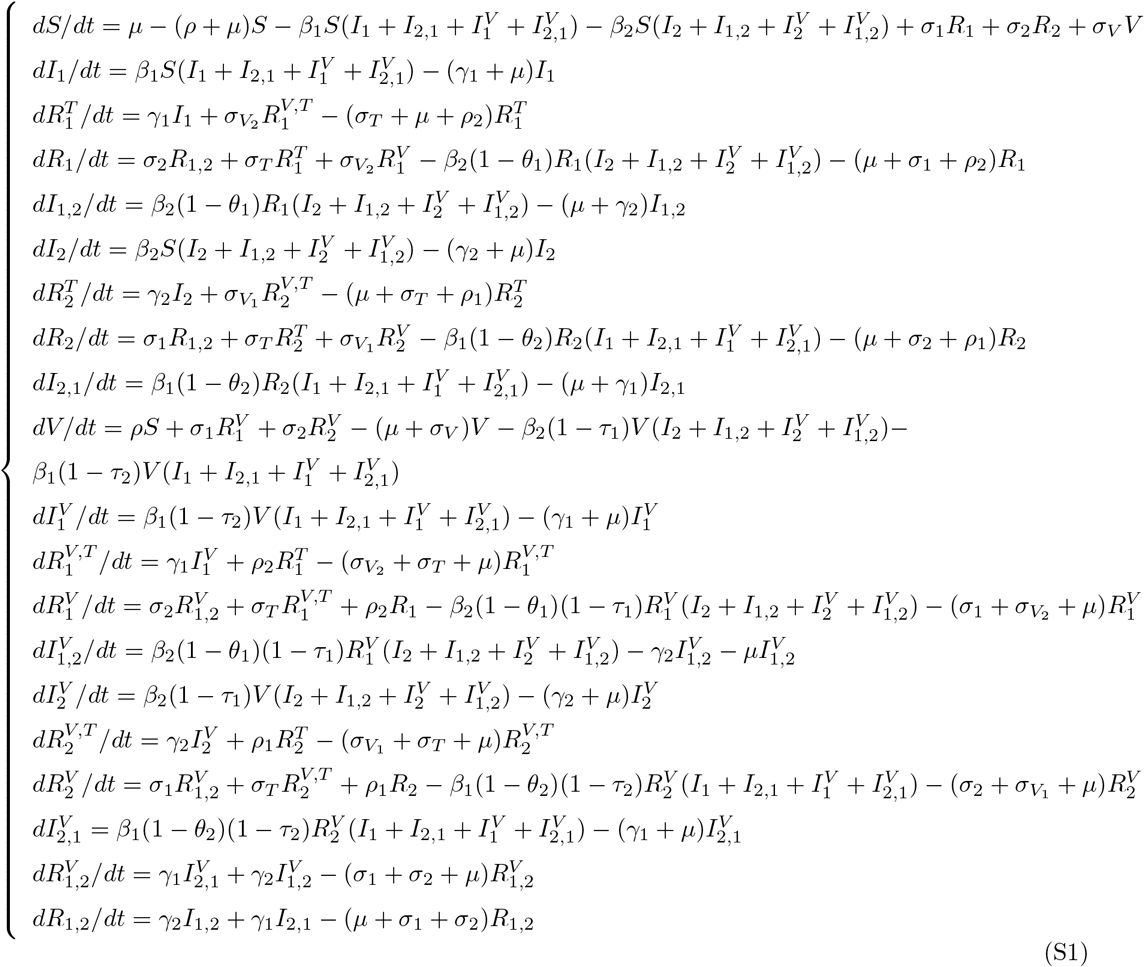

**Table S1:** Compartments in the model.

| Compartment | Definition |
| --- | --- |
| $S$ | Susceptibles to both groups |
| $V$ | Vaccinated |
| $I_i$ | Primary infections by group $i$ |
| $I_{i,j}$ | Secondary infections by group $j$ with a primary infection by group $i$ |
| $I_{i,j}^V$ | Infected individuals by strain $j$, who has been vaccinated (and had group $i$ infection before) |
| $R_{i,j}^T$ | Strain-transcending immune to both strains by primary infection by group $i$ (and a secondary infection by group $j$) |
| $R_{i,j}^V$ | Vaccinated and also immune to group $i$ (and group $j$ from infection) |
| $R_i^{V,T}$ | Vaccinated and also strain-transcending immune to group $i$ by previous infection by group $i$ |
| $R_i$ | Immune to group $i$ |
| $R_{i,j}$ | Immune to both group $i$ and group $j$ |

**Table S2:** Parameters in the model.

| Parameter | Definition | Value | Source of the value |
| --- | --- | --- | --- |
| $R_0^{(i)}$ | Basic reproductive number | 1.44 (H1), 1.60 (H3) | [25] |
| $\beta_i$ | Transmission rate of clade i | $\beta_i(t) = R_0^{(i)}(\gamma_i + \mu)(1 + a \cos(2\pi t))$, $\beta_0 = R_0(\gamma + \mu)$ | / |
| $a$ | amplitude of sinusoidal seasonal forcing function | 0.04 | See methods 2.2 |
| $\gamma_i$ | Recovery rate from infection of clade i | 365/2.64 (H1), 365/3.03 (H3) | [25] |
| $\sigma_T$ | Waning rate of strain-transcending immunity induced by infection | 365/60 | [10] |
| $\sigma_i$ | Waning rate of immunity induced by natural infection of clade i | 1/3.12 (H1), 1/2.28 (H3) | [25] |
| $\theta_i$ | Reduction in susceptibility of the other clade conferred by infection of clade i | / | See methods 2.2 |
| $\sigma_V$ | Waning rate of vaccinal immunity | $\sigma_V = k_2\sigma$ | / |
| $\sigma_{V_i}$ | Waning rate of immunity of the vaccine targeting clade i | $\sigma_{V_i} = k_2\sigma_i$ | / |
| $\rho$ | Vaccination rate | 0-1 | / |
| $\rho_i$ | Vaccination rate of the vaccine targeting clade i | $\rho$ | / |
| $\tau_i$ | Reduction in susceptibility of the other group conferred by vaccination against clade i | $\tau = k_1\theta$ | / |

### S1.3 Invasion analysis of a pandemic variant

We used a similar method as in the previous study [20]. However, our model relaxes the assumption that two strains have the same reproductive ratio and other important epidemiological parameters. We first prepare a population where H3 clade is endemic in the presence of H3 vaccine and H1 clade is absent. We simulate the system for 600 years at semi-weekly time steps to bring H3 to endemic equilibrium, given by

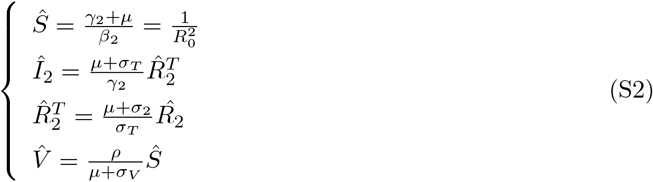

where 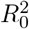 is the basic reproductive ratio of H3. The critical vaccination rate *p*_*c*_ makes the system at disease-free equilibrium, which gives the two following conditions:

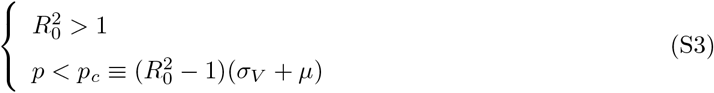

Hence, H3 clade is eliminated when 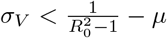 and 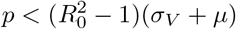.

Next, we introduce a pandemic variant into the population at a very low frequency (*I*_1_ = 10^−6^). The initial invasion dynamics of the pandemic variant approximately follow the equations below.

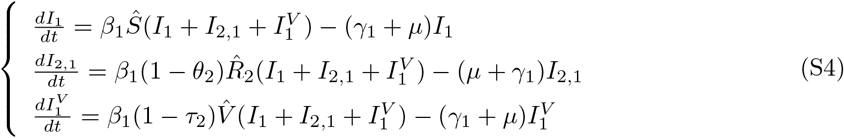

Let 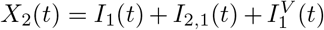 and assume that initially *X*_2_(*t*) approximately grows or decreases exponentially during a short time interval. This exponential behaviour can be written as *Ce*^*xt*^, where *C* is a positive constant and *x* is the invasion rate of the pandemic variant. The equations S4 can be solved as:

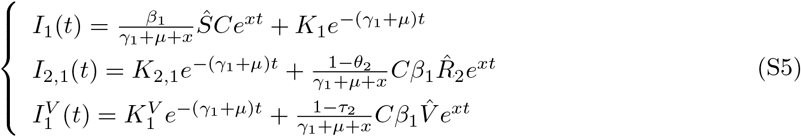

where 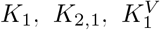 are three constants that depend on initial conditions. If *x > −*(*γ*_1_ + *µ*), then the first term in each function rapidly becomes negligible. Since we initially assumed that 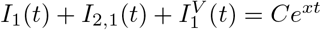 then *x* must satisfy the following relation:

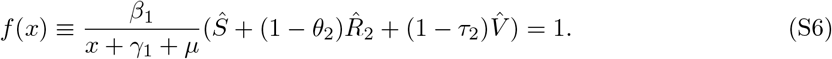

*f* (*x*) is a continuous, decreasing function over (*−*(*γ*_1_ + *µ*), *∞*). If cross-immunity is perfect, i.e. *θ*_2_ = *τ*_2_ = 1, then 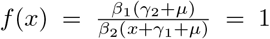, therefore 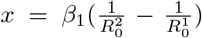 is a solution for (S6). Otherwise, 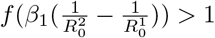, so *f* (*x*) admits a unique solution 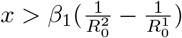 When 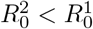, i.e., the basic reproductive ratio of the endemic strain is smaller than the invading variant, *x >* 0, i.e., the pandemic always emerges. Otherwise, 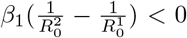, and therefore *x* could be negative or positive. In our case, where *θ*_2_ = 0, the unique solution can be written as below.

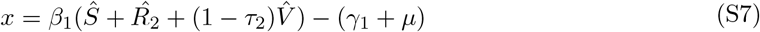

We can see that the vaccine characteristics (*σ*_*V*_, *τ*_2_ and *ρ*) and their relationship with the pathogen epidemiological parameters decide if pandemic emergence could be successful.

### S1.4 Figures

**Figure S1:**
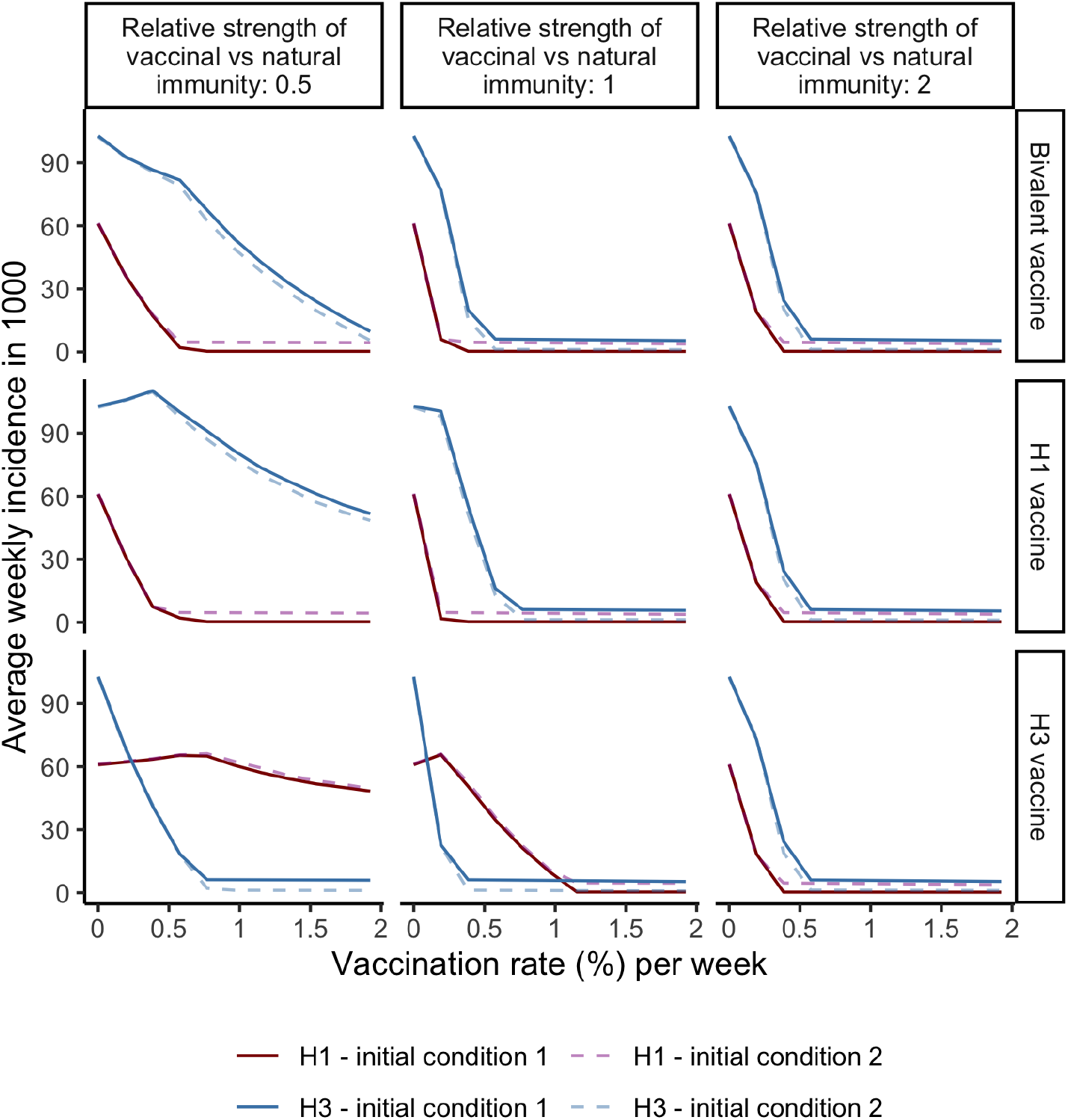
Sensitivity test for initial conditions. The initial condition 1 assumes susceptible population is 89.9%, H1 infection is 0.01% and H3 infection is 0.1%. The initial condition 2 assumes the susceptibles are 89.9%, H1 infection is 0.1% and H3 infection is 0.01%. Average weekly incidence of H1 and H3 subtypes per 1000 population (*y* axis) vaccinated by vaccines with different target clades (rows) and immunity *strengths* (columns, defined as 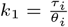), with changes of vaccination rate (%) per week (*x* axis). The results are calibrated with the best-fitting parameters (*θ*_1_ = 0.8, *θ*_2_ = 0.5, *a* = 0.04). Other parameters are the same as in 2.

**Figure S2:**
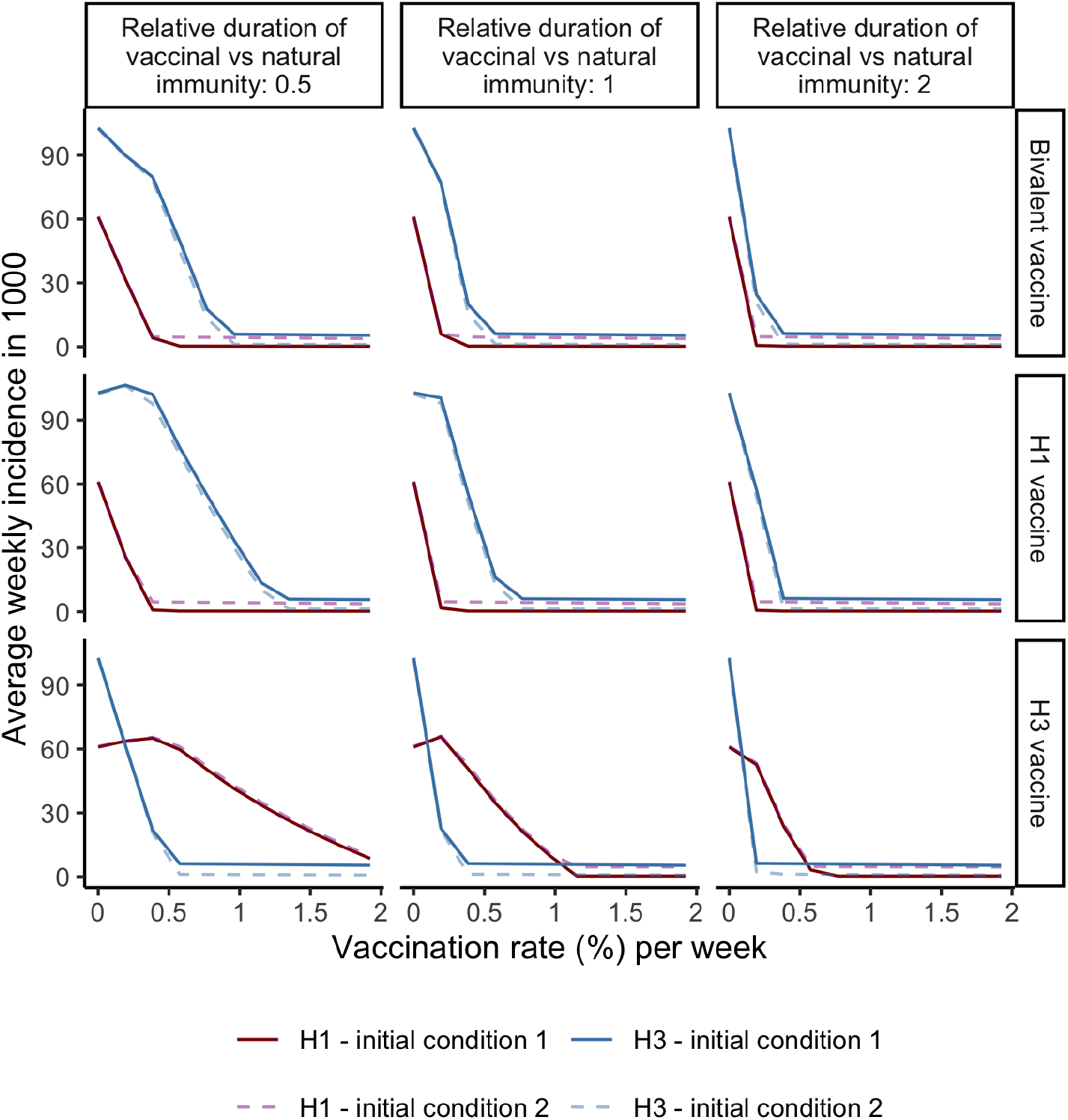
Sensitivity test for initial conditions. Parameters are the same as in S1. Average weekly incidence of H1 and H3 subtypes per 1000 population (*y* axis) vaccinated by vaccines with different target clades (rows) and immunity *durations* (columns, defined as 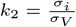), with changes of vaccination rate (%) per week (*x* axis).

**Figure S3:**
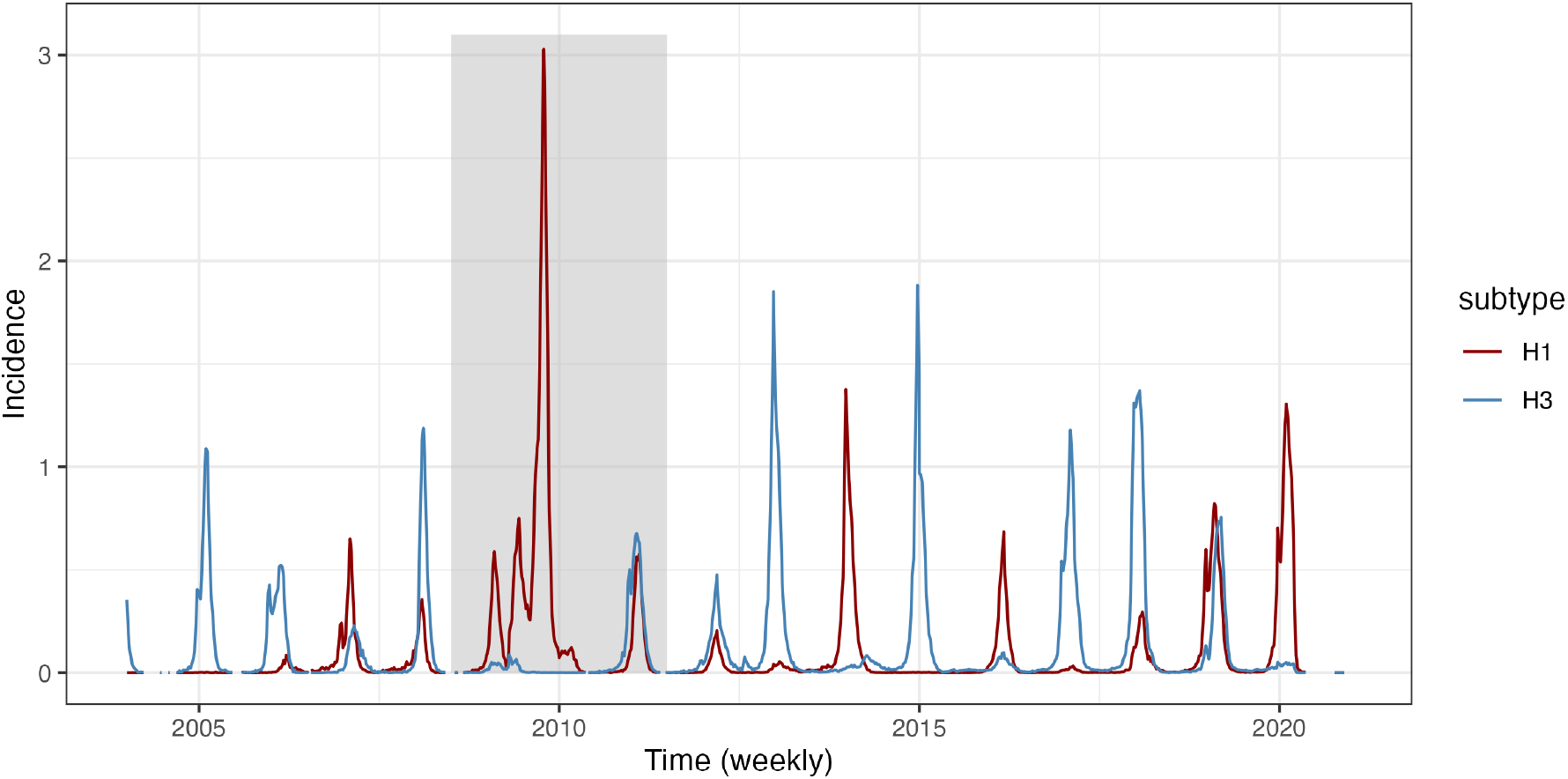
Weekly incidence of human influenza H1N1 (in red) and H3N2 (in blue) in the United States. The grey area is pandemic seasons from 2009-07-01 to 2011-07-01; they are excluded in the model calibration.

**Figure S4:**
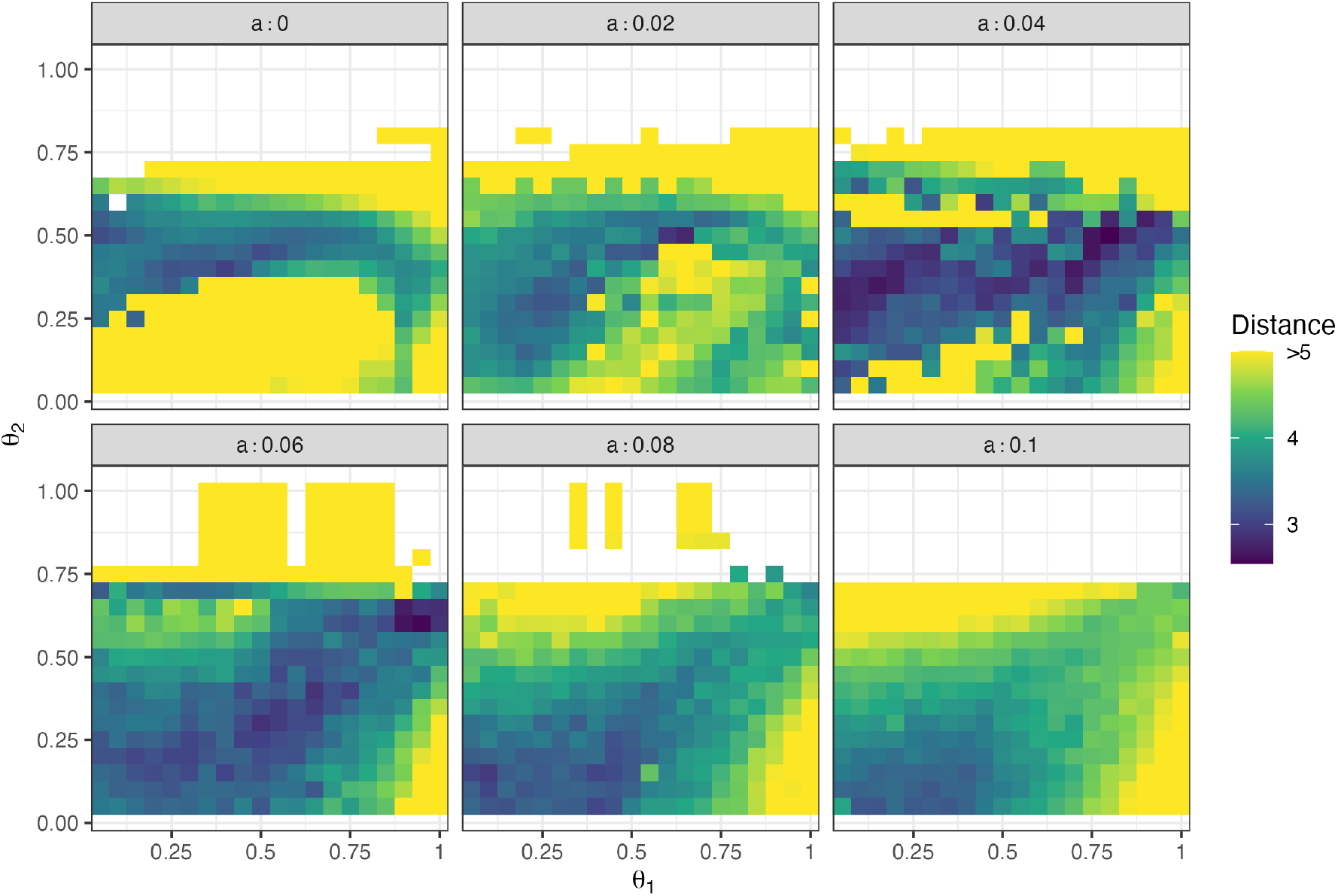
The optimization metric distribution across the parameter space of *a, θ*_1_ and *θ*_2_. Each grid represents a combination of the parameters, the colour of which shows the sum of the absolute distance between simulation and data [11] of three summary statistics, including I) correlation between H1 and H3 seasonal incidence, II) coefficient of variation of H1 (and H3) seasonal incidence, III) auto-correlation of H1 (and H3) seasonal incidence.

**Figure S5:**
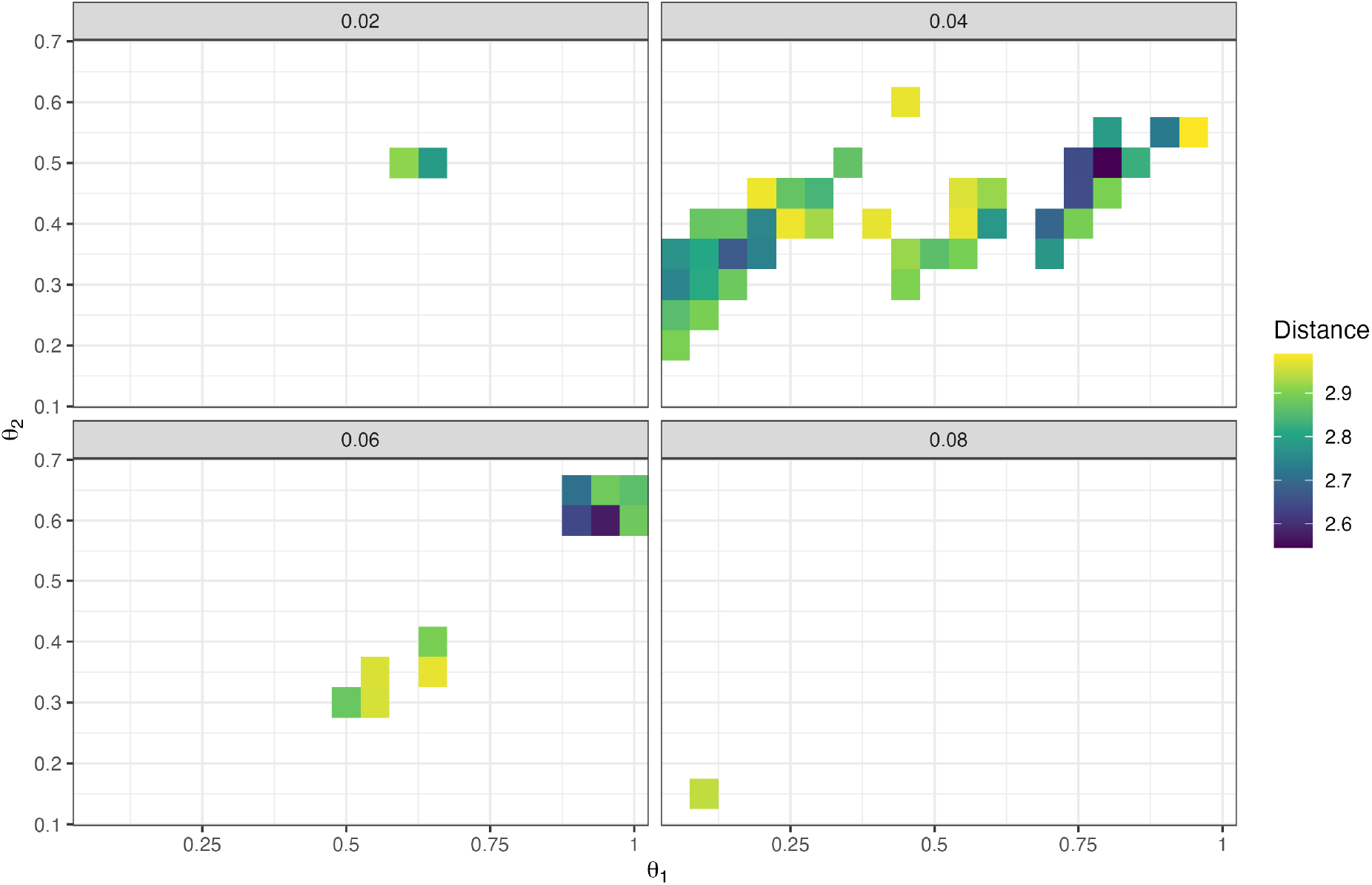
The distribution of the optimization metric (*<* 3) across the parameter space of *a, θ*_1_ and *θ*_2_. Each grid represents a combination of the parameters, the colour of which shows the sum of the absolute distance between simulation and data [11] of three summary statistics, including I) correlation between H1 and H3 seasonal incidence, II) coefficient of variation of H1 (and H3) seasonal incidence, III) auto-correlation of H1 (and H3) seasonal incidence.

**Figure S6:**
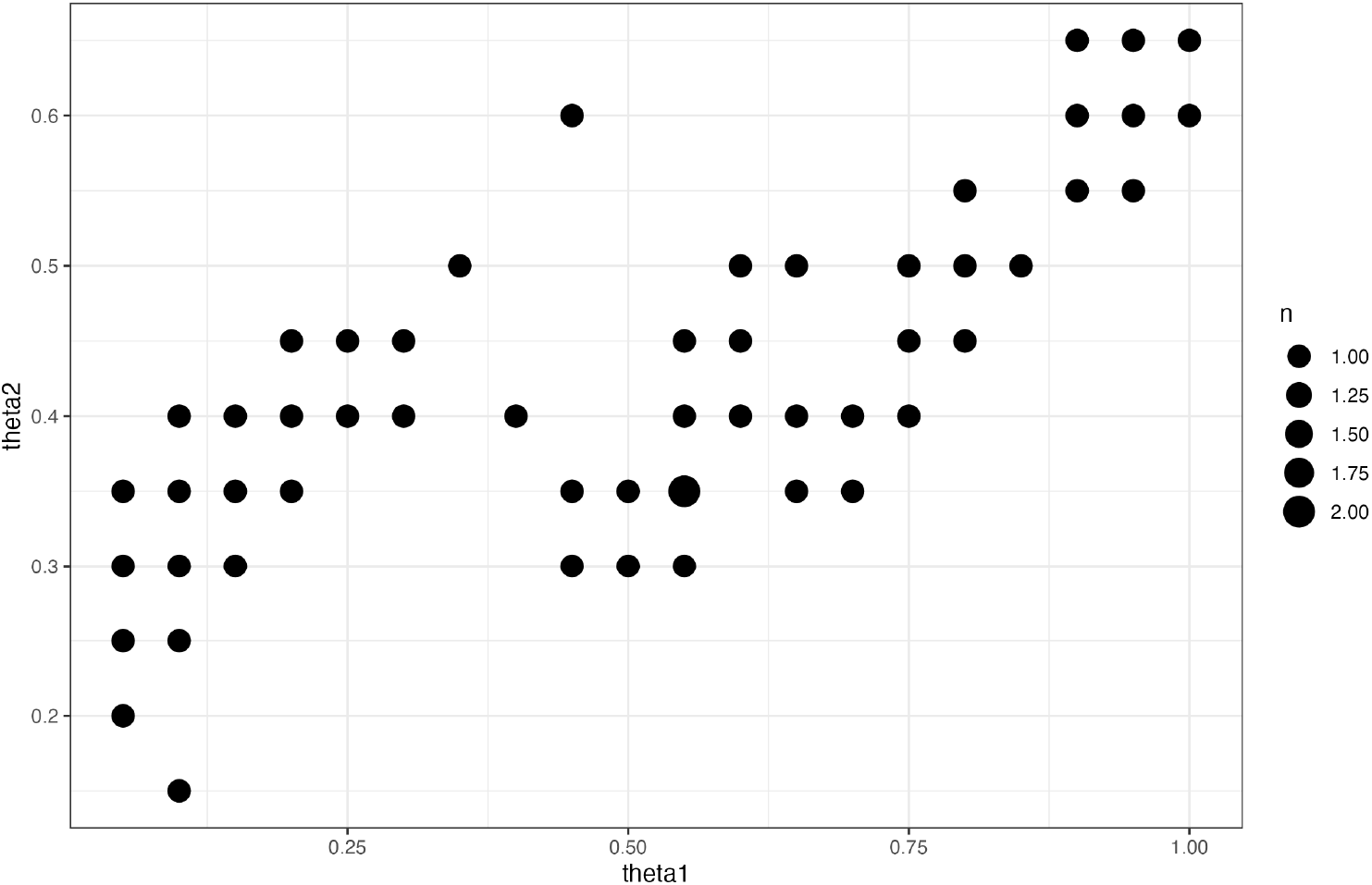
Occurrence counts of the same parameter sets of *θ*_1_ and *θ*_2_ with different *a* that gives optimization metric value smaller than 3.

**Figure S7:**
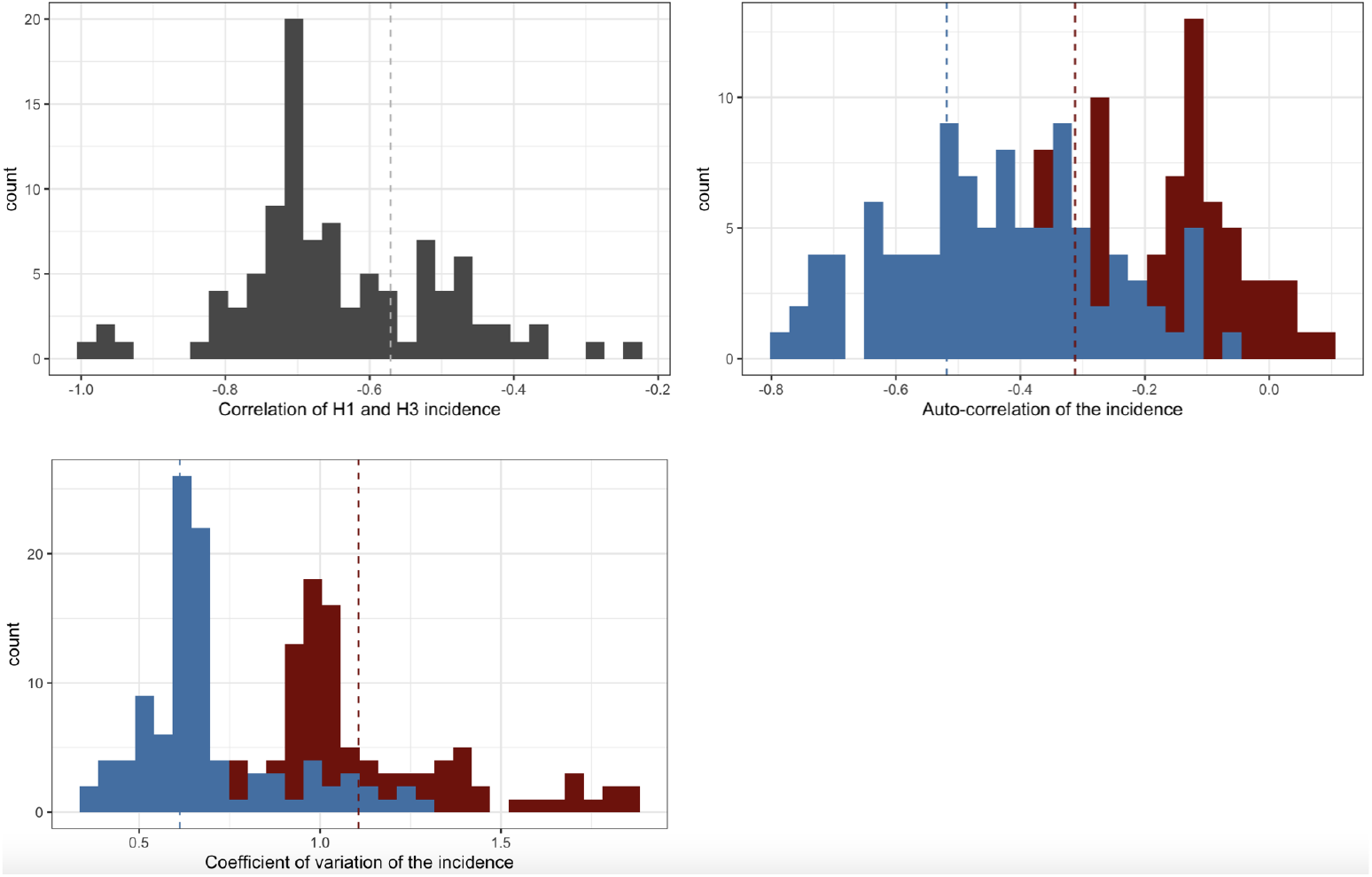
Histogram of correlation between H1 and H3 seasonal incidence (left-top panel), auto-correlation of H1 (and H3) incidence (right-top panel) and coefficient of variation of H1 (and H3) incidence (left-bottom panel) of every 12-yer period of the 2000-year simulation with first 800 years as burn-in, using *θ*_1_ = 0.8, *θ*_2_ = 0.5, *a* = 0.04 as the parameters. Dashed lines show the metric values in the incidence data. Blue and red colours represent H3 and H1 respectively.

**Figure S8:**
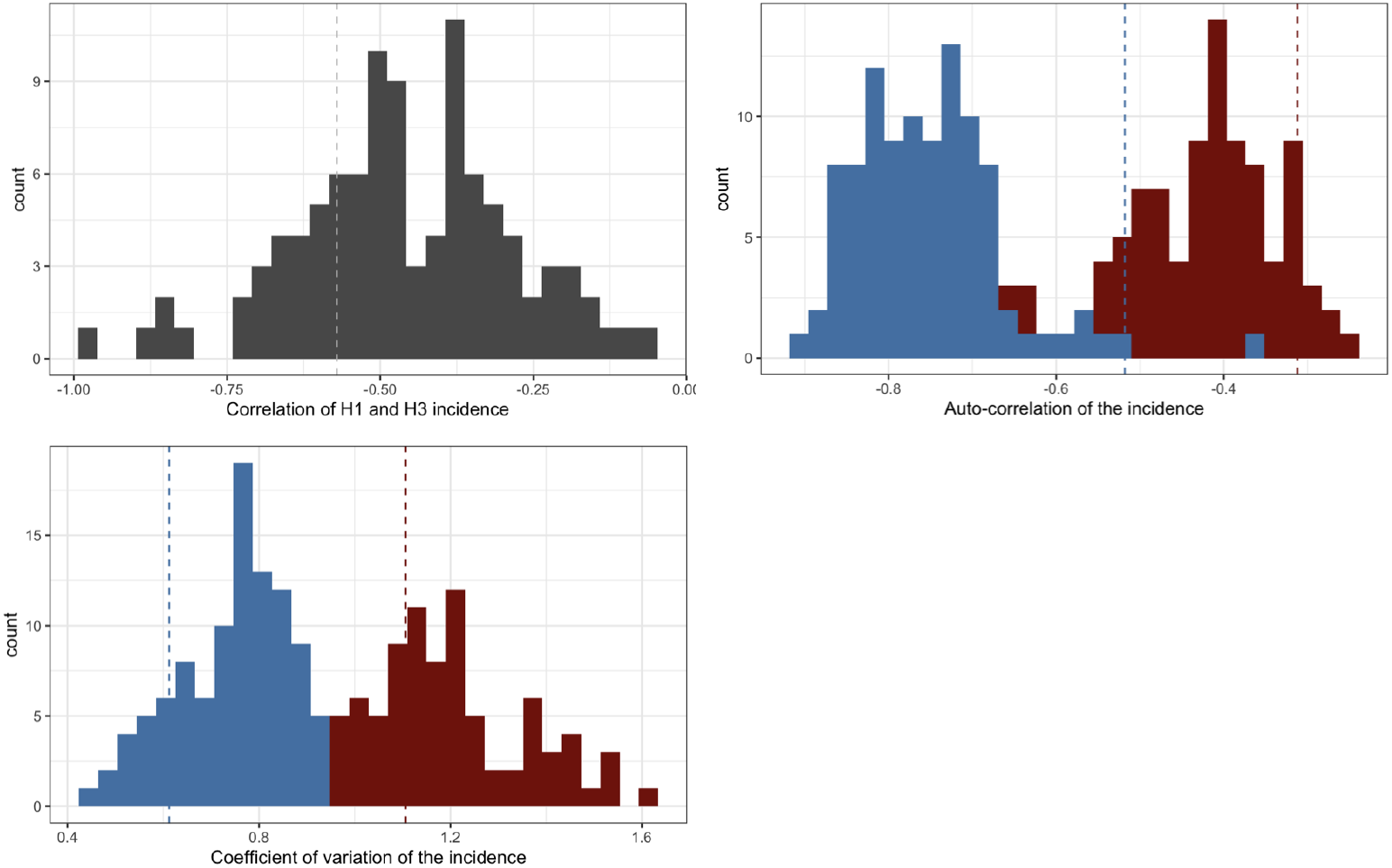
Histogram of correlation between H1 and H3 seasonal incidence (left-top panel), auto-correlation of H1 (and H3) incidence (right-top panel) and coefficient of variation of H1 (and H3) incidence (left-bottom panel) of every 12-yer period of the 2000-year simulation with first 800 years as burn-in, using *θ*_1_ = 0.55, *θ*_2_ = 0.35, *a* = 0.04 as the parameters. Dashed lines show the metric values in the incidence data. Blue and red colours represent H3 and H1 respectively.

**Figure S9:**
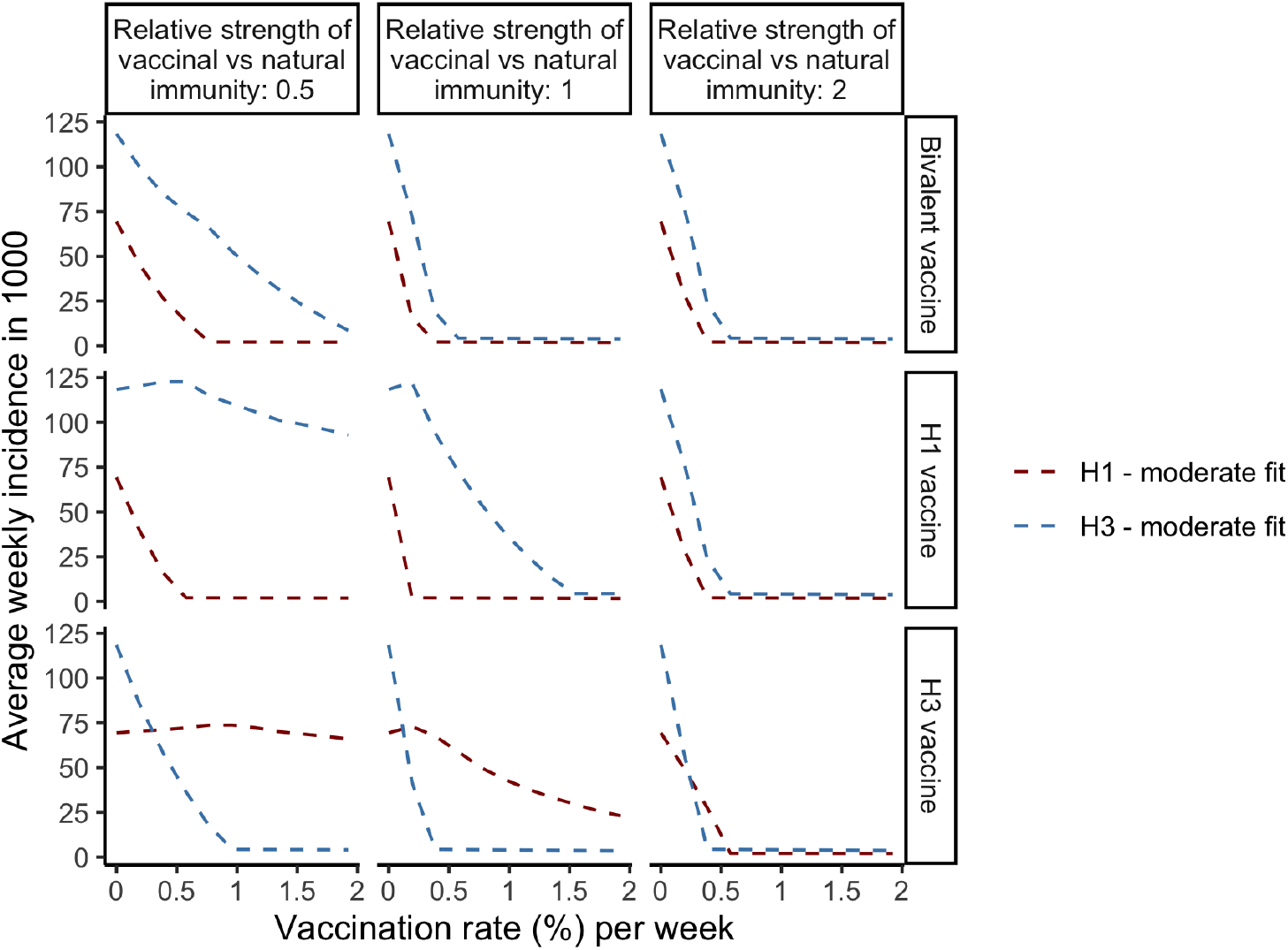
Average weekly incidence of H1 and H3 subtypes per 1000 population (*y* axis) vaccinated by vaccines with different target clades (rows) and immunity *strengths* (columns, defined as 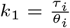), with changes of vaccination rate (%) per week (*x* axis). The results are calibrated with the moderate-fitting parameters (*θ*_1_ = 0.55, *θ*_2_ = 0.35, *a* = 0.04). Other parameters are the same as in 2.

**Figure S10:**
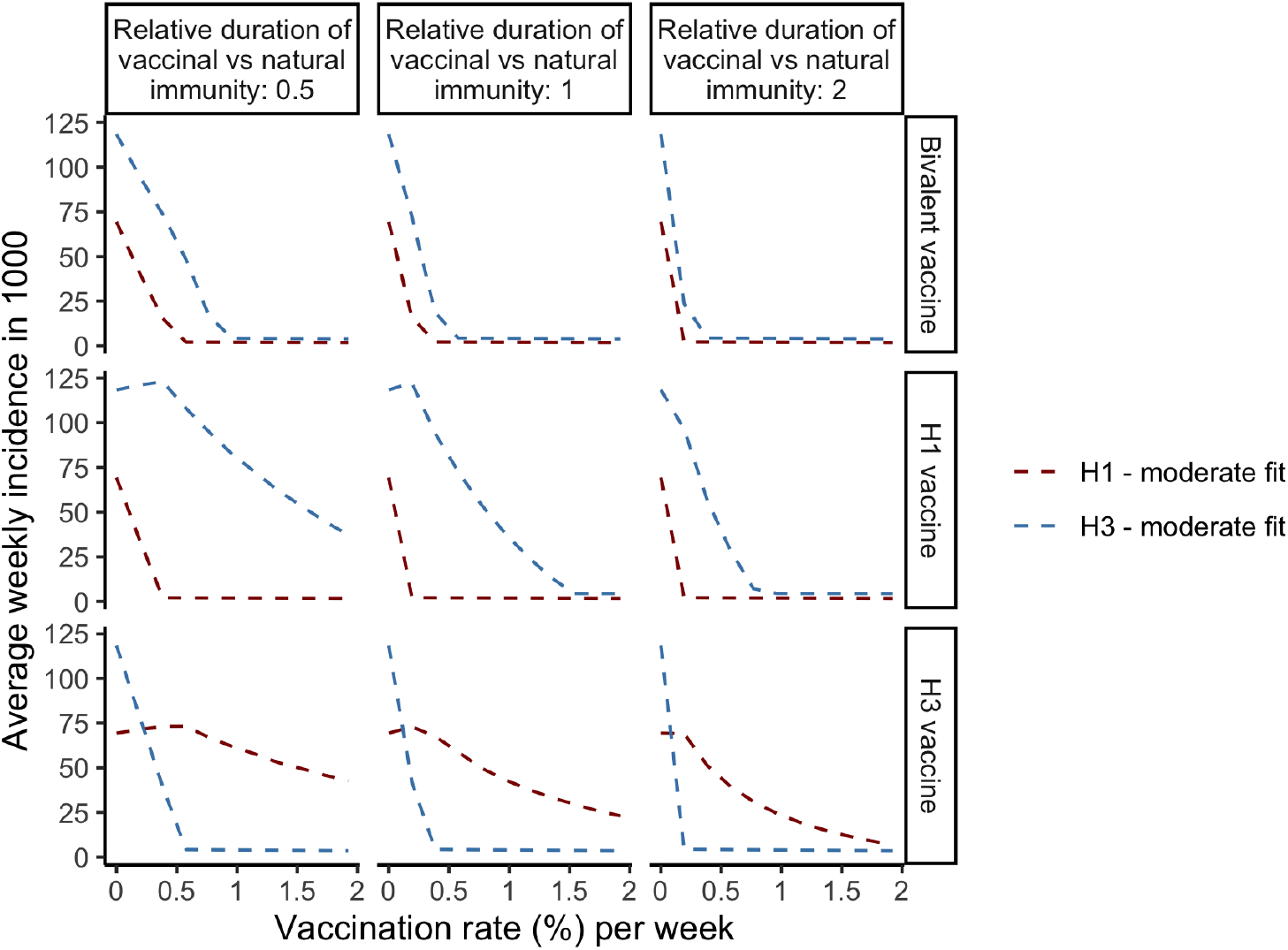
Average weekly incidence of H1 and H3 subtypes per 1000 population (*y* axis) vaccinated by vaccines with different target clades (rows) and immunity *durations* (columns, defined as 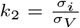), with changes of vaccination rate (%) per week (*x* axis). The parameters are the same as in S9.

**Figure S11:**
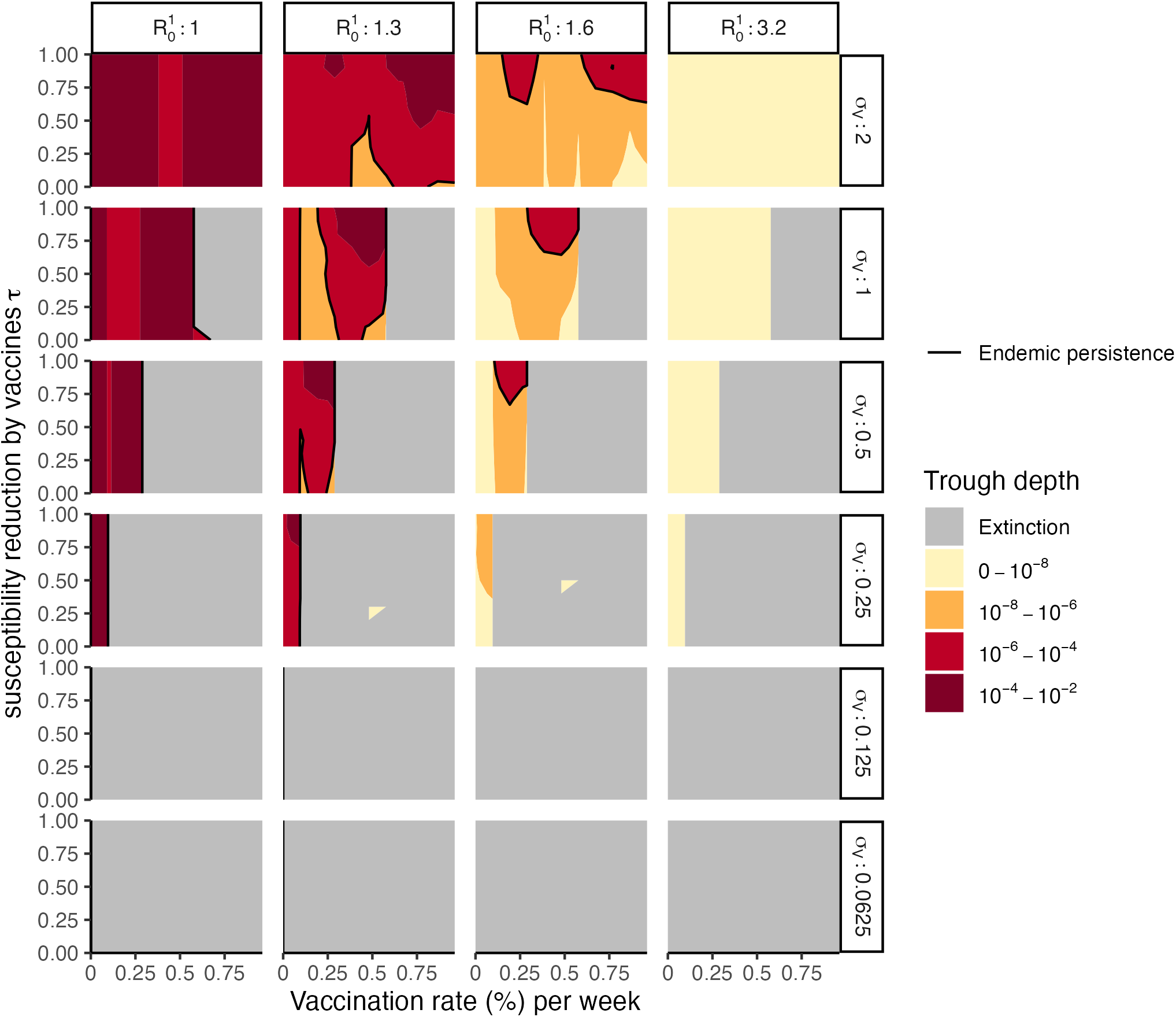
Persistence of the endemic variant 5 years after its emergence, measured by trough depth of its incidence ((the minimal incidence), under vaccination of different immunity strength (*y* axis), duration (row) and vaccination rate *ρ* (%) per week (*x* axis), with different basic reproduction number 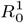 (column). The grey area is when the endemic strain fails to persist. The black curve indicate the transition between persistence and failure in the persistence of the endemic strain. Other parameters are same as those in Figure 4.

**Figure S12:**
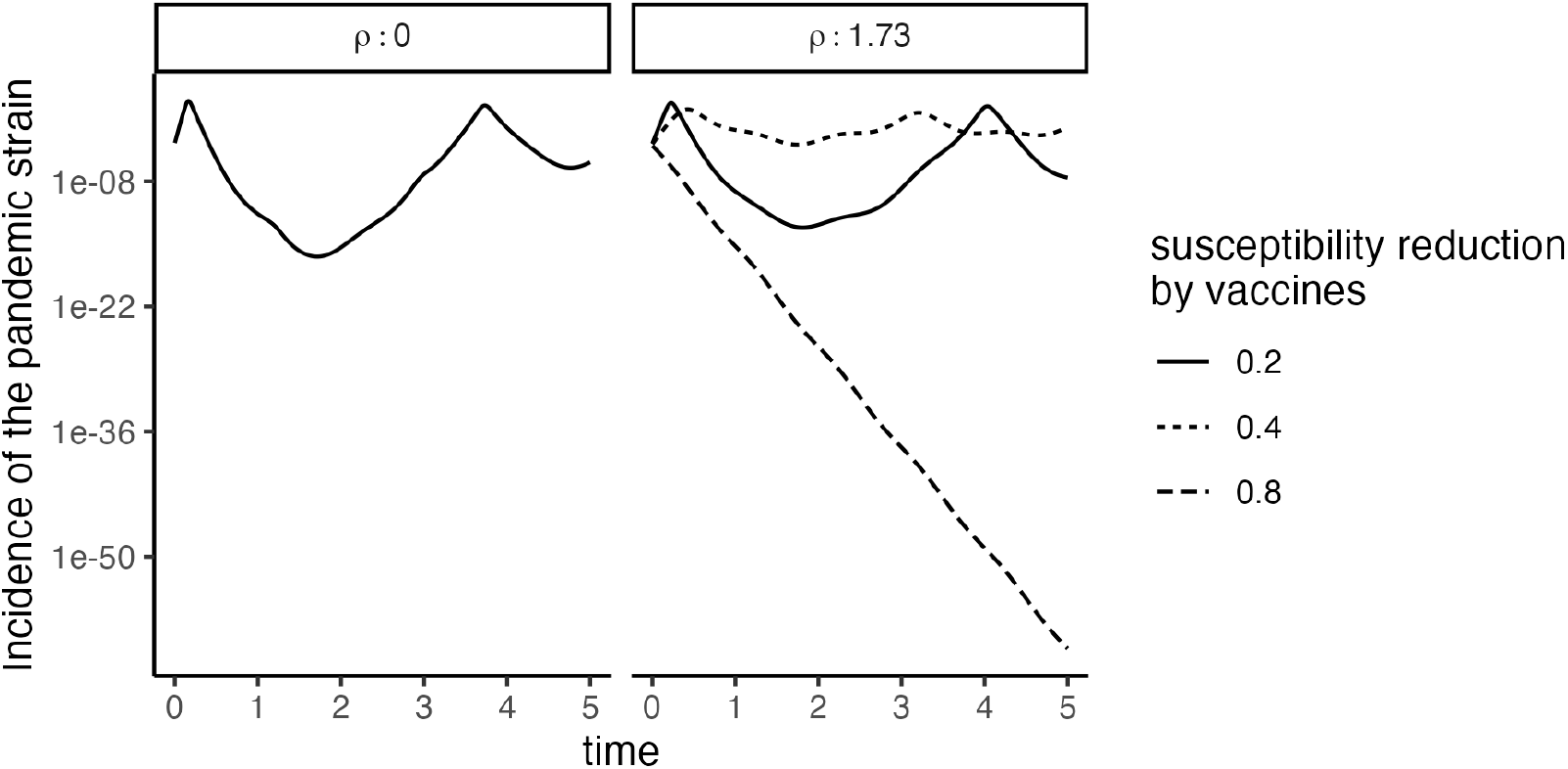
Deterministic trajectories after the pandemic strain (at the incidence of 10^−6^) invades the endemic equilibrium of H3. Line types represent weak (20%), middle (40%), and strong (80%) susceptibility reduction against the pandemic strain (*τ*_2_) by the vaccine.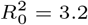, 1/*σ*_*V*_ = 16(*yr*), *ρ* = 1.73% per week. Other parameters are as in Figure 4.

